# High blood viscosity in acute ischemic stroke

**DOI:** 10.1101/2023.06.22.23291757

**Authors:** Jihoon Kang, Ju Seok Oh, Beom Joon Kim, Jun Yup Kim, Do Yeon Kim, So-Yeon Yun, Moon-Ku Han, Hee-Joon Bae, Inwon Park, Jae Hyuk Lee, You Hwan Jo, Kyung Hyun Ahn

## Abstract

**Background:** Changes in blood viscosity can influence the shear stress level at the vessel wall. However, there is limited evidence to date regarding the role of high blood viscosity in acute thrombogenic events. We aimed to investigate the effect of blood viscosity on stroke occurrence and the clinical utility of blood viscosity measurements obtained immediately upon hospital arrival.

**Methods:** Patients with suspected stroke visiting the hospital within 24 h of the last known well time were enrolled. Point-of-care testing was used to obtain blood viscosity measurements before intravenous fluid infusion. Blood viscosity was measured as the reactive torque generated at three different oscillatory frequencies (1, 5, and 10 *Hz*). Blood viscosity results were compared among patients with stroke mimics, ischemic stroke, and hemorrhagic stroke.

**Results:** Among 112 enrolled patients, blood viscosity measurements were accomplished within 2.4 ± 1.3 min of vessel puncture. At an oscillatory frequency of 10 *Hz*, blood viscosity differed significantly between the ischemic stroke (24.2 ± 4.9 *cP*) and stroke mimic groups (17.8 ± 6.5 *cP*, P < 0.001). This finding was consistent at different oscillatory frequencies (134.2 ± 46.3 vs. 102.4 ± 47.2 at 1 *Hz* and 39.2 ± 11.5 vs. 30.4 ± 12.4 at 5 *Hz*, Ps < 0.001), suggesting a relationship between decreases in viscosity and shear rate. Among patients with ischemic, blood viscosity values were lower (16.4 ± 3.3) in those who had received intravenous fluid before blood sampling than in those who had not. The area under the receiver operating curve for differentiating cases of stroke from stroke mimic was 0.79 (95% confidence interval, 0.69 – 0.88).

**Conclusion:** Patients with ischemic stroke exhibit increases in whole blood viscosity when admitted within 24 h of last known well time, suggesting that blood viscosity measurements can aid in differentiating ischemic stroke from other diseases.

## Introduction

Hemostatic balance is essential for maintaining normal physiological conditions, especially even in patients with risk factors such as atrial fibrillation, high blood pressure, and rupture-prone atherosclerosis.^1^ A sudden breakdown event of hemostasis provokes a transition to a prothrombotic state of activating coagulation pathways and gathering the prothrombotic molecules, which leads to thrombus formation, an inexorable pathologic stage, such as the development of ischemic stroke.^1, 2^

Endothelial damage is a pivotal step in initiating thrombotic events.^3^ The blood circulates consistently by imposing a force on the vascular wall; however, when this force exceeds a threshold, damage to the endothelium or pre-existing plaque rupture can occur, promoting subsequent pro-coagulation processes.^4, 5^ In terms of hemodynamics, the force by blood flow is characterized by shear stress, defined as the product of shear rate and blood viscosity. The shear rate is measured at 1,000 *s^−1^* in normal arteries, although this rate can increase to over 5,000 *s^−1^* in some forms of pathologic atherosclerosis.^6^ Such high shear rates exacerbate the progression of atherosclerosis and substantially increase the risk of rupture.^7, 8^

However, given the long-term stability of the shear rate effect, it alone cannot adequately explain the rapid differential change of physical shear stress in the vasculature. In addition, it needs to draw attention to the blood viscosity, another determinant of the shear stress level, which may also account for fluctuating conditions caused by the complex interactions among heterogeneous constituents involved in various biological processes.^9, 10^ However, there is limited evidence to date regarding the role of high blood viscosity in acute thrombogenic events.^11, 12^

In the present study, we aimed to investigate the effect of blood viscosity on stroke occurrence and the clinical utility of blood viscosity measurements obtained immediately upon hospital arrival. To achieve this aim, we compared blood viscosity measurements between patients with suspected stroke, patients with non-thrombogenic disease, and healthy controls using a parallel plate rheometer designed for point-of-care testing (POCT), allowing real-time measurement after blood sampling.^13^ This method is particularly advantageous in ensuring more accurate measurements of pure blood viscosity because it can avoid using any additional anticoagulant such as ethylenediaminetetraacetic acid.^11, 14^

## Methods

### Study population and data collection

Patients who visited the emergency department with focal neurological symptoms at least 24 h after the last known well time were enrolled. They were triaged with suspected stroke at the time of arrival and underwent rapid neurological assessment, laboratory tests, and imaging studies in accordance with the institution’s clinical pathway protocol (**Supplemental Figure 1**). Informed consent for study participation and additional blood samples were obtained within 15 min of hospital arrival. Blood sampling was performed prior to the administration of intravenous fluid or medication to avoid the influence of external factors on viscosity measurements. Cases in which blood sampling was deficient or delayed due to vascular fragility or low blood volume were excluded.

For all enrolled patients, diagnoses were confirmed by blinded neurologists based on medical information and imaging studies after discharge, following which patients were classified into ischemic stroke, hemorrhagic stroke, and stroke mimic (or non-stroke) groups. In some cases, patients transferred due to ischemic stroke from other institutions were separately classified into an ischemic stroke with intravenous (IV) fluid group. The following clinical data were also collected for all patients: demographic information; cardiovascular risk factors including hypertension, diabetes mellitus (DM), and dyslipidemia; initial laboratory findings including white blood cell (WBC) count, hemoglobin, hematocrit, platelets, lipid battery results, C-reactive protein (CRP), D-dimer, fibrinogen, glucose, blood urea nitrogen (BUN), creatinine, and prothrombin time (PT).

### Study protocol and consent

The local institutional review board of Seoul National University Bundang Hospital (Republic of Korea) approved this study. Patients or caregivers provided written informed consent for additional blood sampling and viscosity measurements. The use of registered data is permitted upon request after the review.

### Estimation of blood viscosity

Blood viscosity was measured using a parallel plate rheometer (ARS-Medi, Advanced Rheology Solutions, Ltd., Republic of Korea) designed for point-of-care and approved by the Korean Regional Food and Drug Administration. This method has been widely adopted for micro-scale structures and dispersed fluids as a fast and accurate measurement without data loss (**Supplemental Table 1**).^15, 16^ A 2-*mL* whole blood sample was evenly distributed on the parallel plate (**Figure 1**). Testing was performed within 3 min of sampling and without anticoagulant reagents to ensure measurement of pure blood values. Oscillation frequencies were 1, 5, and 10 *Hz*, and the resultant values at each frequency were used to estimate the reactive torque from the blood.

**Figure 1.**
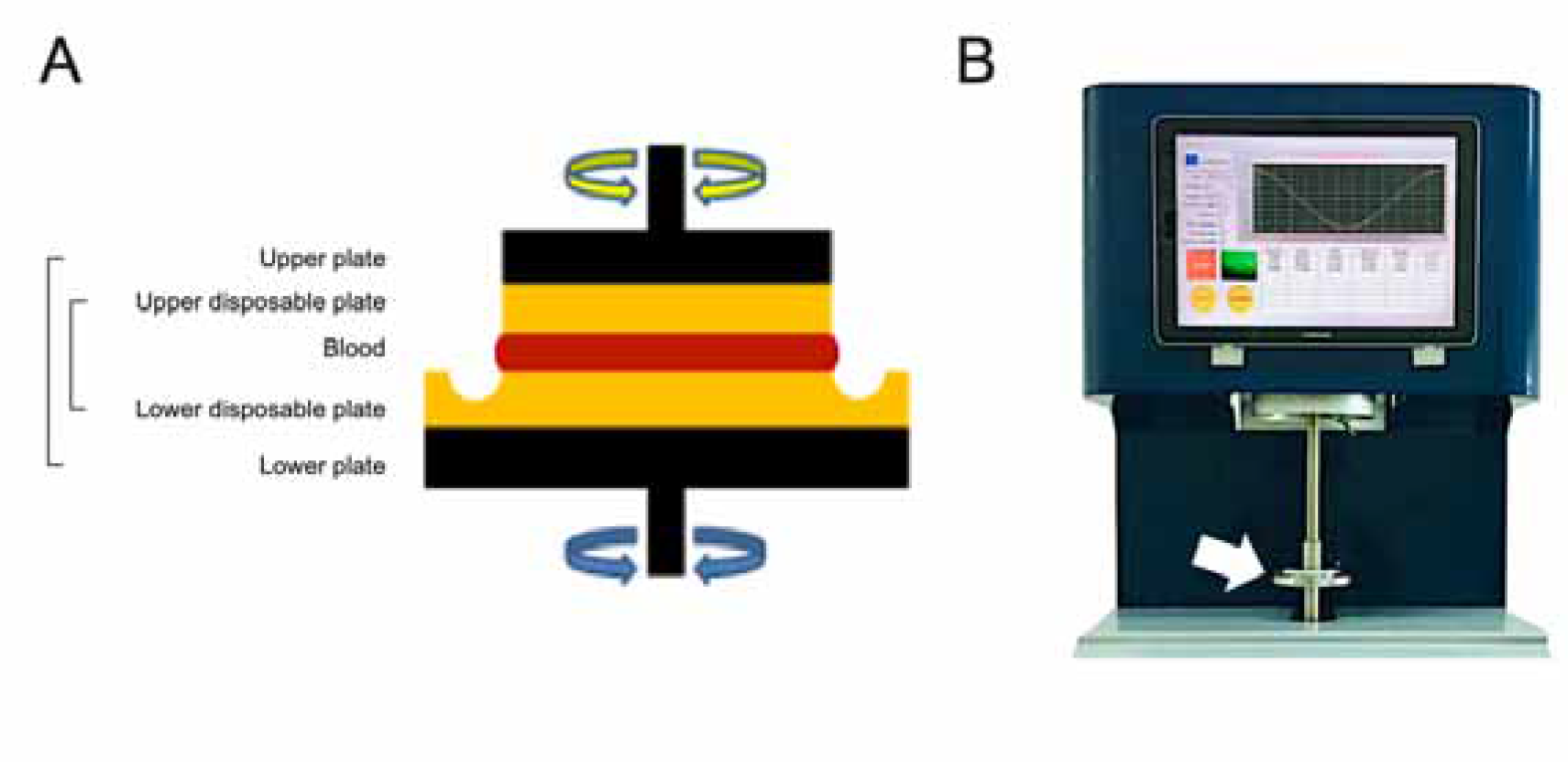
Schematic illustration of the core part measuring the blood viscosity of parallel plate rheometer (A) and the overall feature of ARS-Medi device (B). After raising the upper metal plate (black color), a disposable plate was installed (yellow color). Blood samples were then infused into the disposable plate, and the upper metal plate was lowered in increments of 5 *mm* to adjust the rotational oscillatory frequency (A). The device measured the reactive torque generated by the blood to estimate the blood viscosity.

### Statistical analysis

Baseline characteristics, blood laboratory profiles, and blood viscosity of study population were summarized. Correlations between blood viscosity and various laboratory profiles were analyzed using the Pearson correlation test.

The blood viscosity among diagnoses of ischemic stroke, ischemic stroke with IV fluid, hemorrhagic stroke, and stroke mimic were compared by the analysis of variance (ANOVA) test. Then, the ability of blood viscosity to differentiate between ischemic stroke and stroke mimics was tested using the area under the receiver operating curve (AUROC). All statistical analyses were conducted using SPSS (version 22.0, IBM) and R software (version 4.2.1).

## Results

It enrolled 112 patients with a mean age of 69.1 ± 13.3 years, 56.9% of whom were male, after excluding those with low blood sample volume (< 1 *mL*) and time delay during sampling. POCT measurements successfully obtained the blood viscosity within an average of 2.4 ± 1.3 min following a venular puncture. Blood viscosity values decreased with increasing the oscillation frequency, which was 115.4 ± 47.8 *cP*, 34.2 ± 12.2 *cP*, and 20.3 ± 6.5 *cP* at oscillatory frequencies of 1, 5, and 10 *Hz*, respectively (**Supplemental Figure 2**). Blood viscosity exhibited significant correlations with hemoglobin (*γ* = 0.27, P by Pearson correlation test = 0.004) but not with WBC count (*γ* = 0.10, P = 0.52), hematocrit (*γ*= 0.23, P = 0.08). Among blood chemistry tests, total cholesterol and BUN were significantly correlated with blood viscosity (*γ* = 0.26, P = 0.01 and *γ* = −0.21, P=0.03, **Supplemental Table 2**).

### Disease groups and blood viscosity

The stroke mimics coalesced by various diseases comprised 45.3%, and ischemic stroke (35.9%) and hemorrhagic stroke (5.4%) were followed. There were significant differences in the proportion of dyslipidemia and values of CRP and INR levels according to the disease groups (**Table 1**). The higher CRP values of the stroke mimic group were observed in systemic infection or encephalopathy (**Supplemental Table 3**).

**Table 1.** Baseline characteristics and diagnoses of enrolled patients (n = 112)

| Variables | Stroke mimic<br>(n = 53) | Ischemic stroke<br>(n = 53) | Hemorrhagic stroke<br>(n = 6) | P |
| --- | --- | --- | --- | --- |
| Age, mean ± SD | 67.1 ± 14.2 | 70.0 ± 12.4 | 70.0 ± 13.8 | 0.44 |
| Male | 28 (52.8%) | 36 (67.9%) | 1 (16.7%) | 0.03 |
| Hypertension | 23 (43.4%) | 31 (58.5%) | 2 (33.3%) | 0.21 |
| Diabetes mellitus | 15 (28.3%) | 17 (32.1%) | 2 (33.3%) | 0.90 |
| Dyslipidemia | 6 (11.3%) | 26 (49.1%) | 1 (16.7%) | < 0.001 |
| WBC (103/μL) | 9.2 ± 6.6 | 6.9 ± 2.0 | 13.1 ± 11.6 | 0.93 |
| Hemoglobin (g/dL) | 13.3 ± 2.5 | 14.0 ± 2.0 | 14.4 ± 1.9 | 0.18 |
| Hematocrit (%) | 39.8 ± 6.9 | 42.1 ± 5.4 | 42.8 ± 5.3 | 0.12 |
| Platelet (103/μL) | 224.7 ± 84.1 | 224.0 ± 64.6 | 265.3 ± 102.0 | 0.45 |
| Fibrinogen (mg/dL) | 392.6 ± 162.2 | 325.2 ± 72.7 | 330.5 ± 122.2 | 0.03 |
| D-dimer (g/L) | 1.7 ± 1.6 | 0.9 ± 1.1 | 0.5 ± 0.01 | 0.22 |
| CRP (mg/L) | 3.8 ± 7.7 | 0.5 ± 0.8 | 1.1 ± 1.1 | 0.03 |
| Glucose (mg/dL) | 132.9 ± 51.9 | 131.3 ± 42.7 | 138.3 ± 39.2 | 0.90 |
| Total cholesterol<br>(mg/dL) | 159.9 ± 47.7 | 174.6 ± 45.4 | 197.5 ± 76.3 | 0.17 |
| HDL (mg/dL) | 49.2 ± 16.0 | 50.1 ± 11.9 | 62.7 ± 22.2 | 0.21 |
| LDL (mg/dL) | 91.7 ± 33.1 | 107.4 ± 40.9 | 121.7 ± 54.4 | 0.10 |
| BUN (mg/dL) | 26.2 ± 29.4 | 17.0 ± 6.7 | 23.2 ± 15.4 | 0.17 |
| Creatinine (mg/dL) | 2.8 ± 1.2 | 0.9 ± 0.3 | 1.0 ± 1.0 | 0.72 |
| INR | 1.0 ± 0.1 | 1.0 ± 0.1 | 1.5 ± 1.2 | < 0.001 |

Values represent the number of patients (percentage) or mean ± SD as appropriate. P values were obtained using the Pearson $\chi^2$ test and analyses of variance, as appropriate.

Blood viscosity values were plotted with the disease groups (**Figure 2**). At an oscillatory frequency of 10 *Hz*, blood viscosity was 24.2 ± 4.9 *cP* in patients with ischemic stroke, which was significantly different from 17.8 ± 6.5 *cP* with stroke mimic, 23.2 ± 5.9 *cP* with hemorrhagic stroke (*P* by *ANOVA* < 0.001). The significant associations of higher viscosity values of ischemic stroke were consistently observed at lower oscillatory frequencies of 1 *Hz* (134.2 ± 46.3 vs. 102.4 ± 47.2 *cP*, *P* = 0.001) and 5 *Hz* (39.2 ± 11.5 vs. 30.4 ± 12.4 *cP*, *P* = 0.003, **Figure 3 and Supplemental Table 4)**. Among patients diagnosed with ischemic stroke, blood viscosity at 10 Hz oscillatory frequency in those who received IV fluid before blood sampling was 16.4 ± 3.3 cP, which was similar to that observed in the stroke mimic group. The discrimination performance of blood viscosity measurements in the emergency setting exhibited good performance among the disease groups (AUROC = 0.79, **Figure 4**).

**Figure 2.**
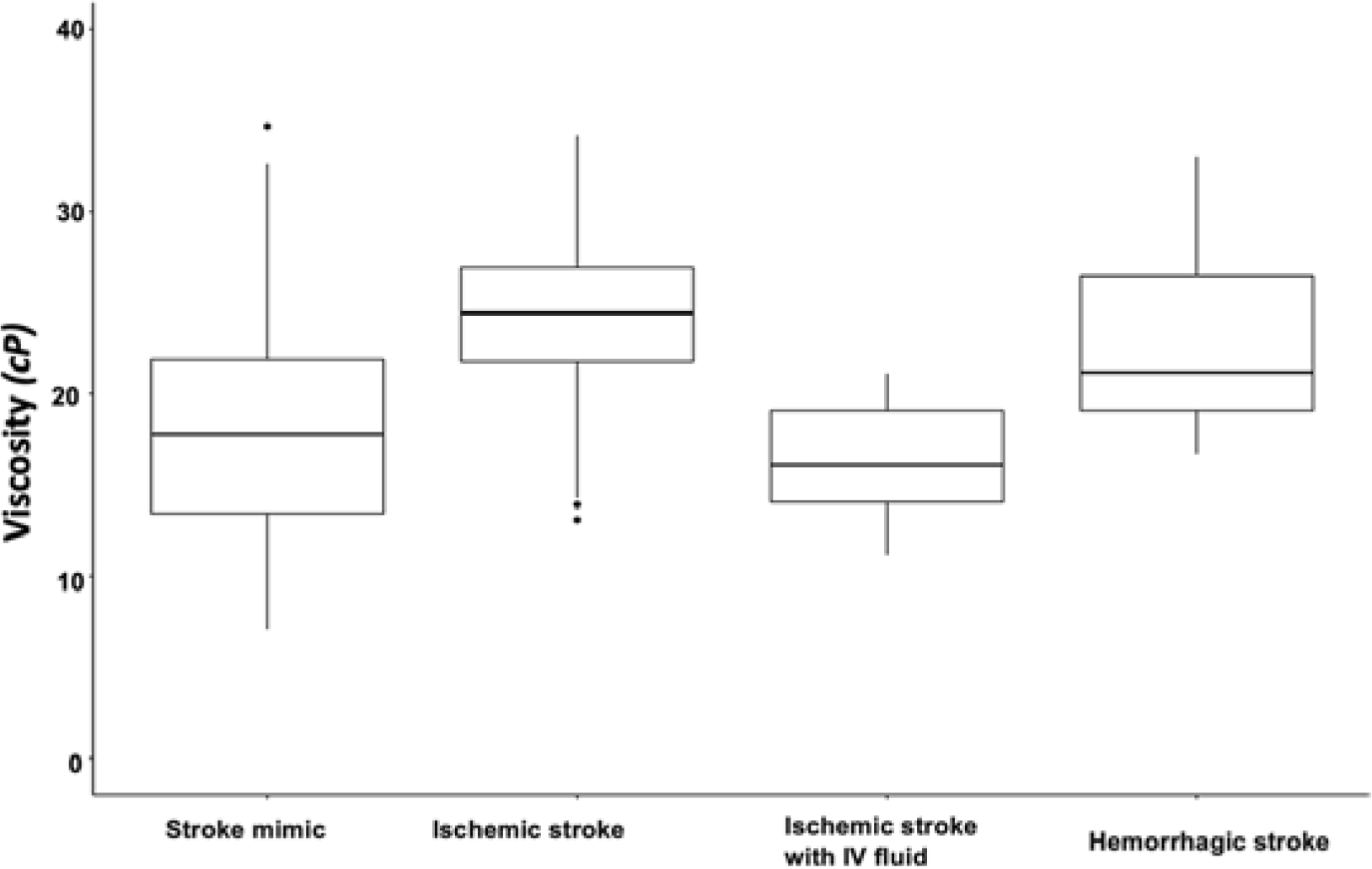
Box plot of blood viscosity according to disease groups. The graph shows the blood viscosity distribution in each disease group at the oscillatory frequency of 10 *Hz* (midline within the box for median values and the box itself for interquartile range). The blood viscosity value was 24.2 ± 4.9 *cP* in patients with ischemic stroke, which were significantly different from the stroke mimics (17.8 ± 6.5 *cP*), hemorrhagic stroke (23.2 ± 5.9 *cP*), and ischemic stroke with intravenous fluid infusion before the measurement (16.4 ± 3.3 *cP*).

**Figure 3.**
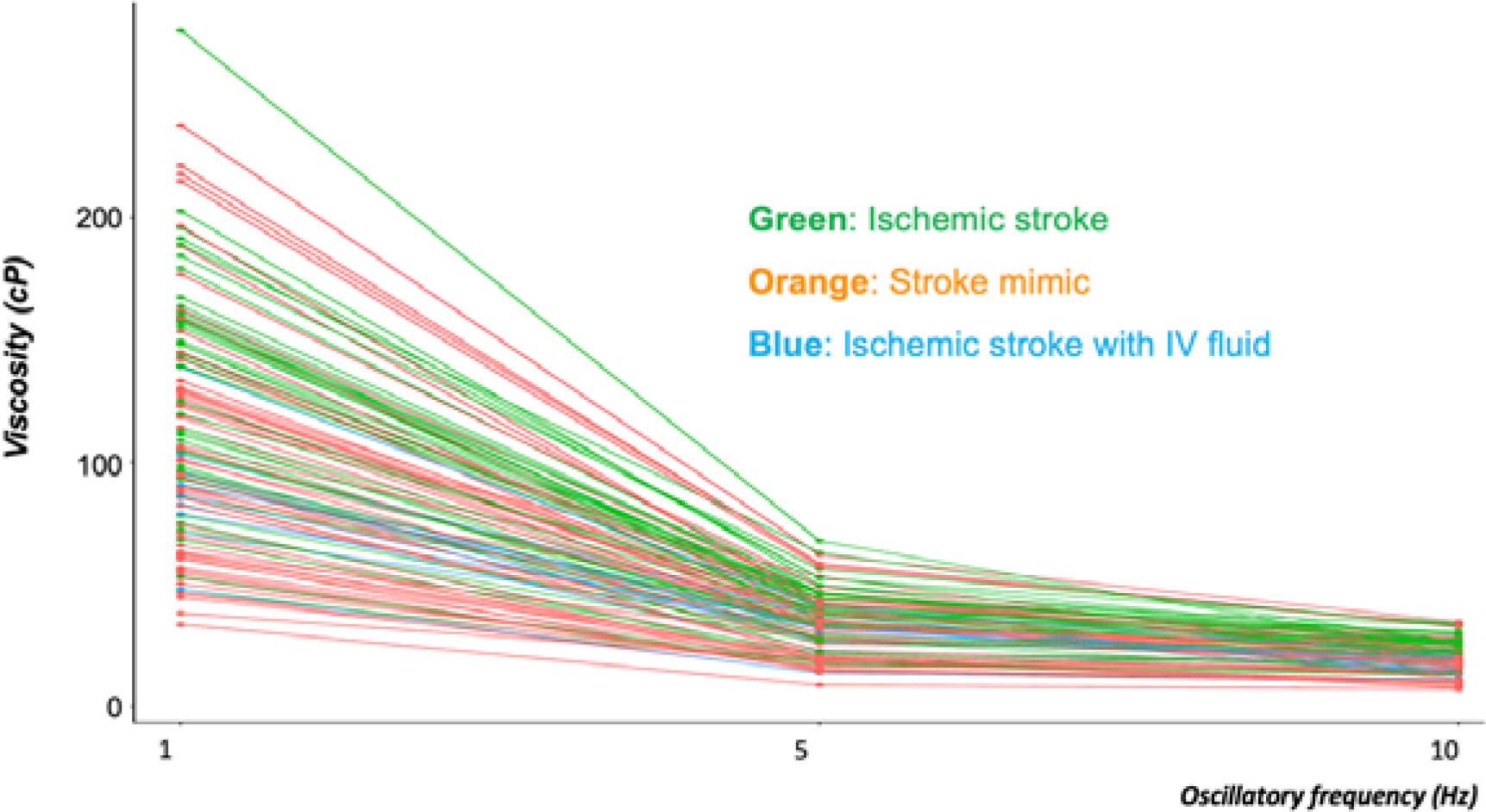
Distribution of blood viscosities in the ischemic stroke, ischemic stroke with IV fluid, and stroke mimic groups at three different oscillatory frequencies. The graph showed every inter-personal viscosity value (Y-axis) across the different oscillatory frequencies (X-axis). The line connected the individual measurement values. At each of the three oscillatory frequencies, blood viscosity was significantly higher in patients with ischemic stroke (green line) than in those with stroke mimic (orange line). Decreases in blood viscosity were observed in patients with ischemic stroke who had been treated with intravenous (IV) fluid (blue line).

**Figure 4.**
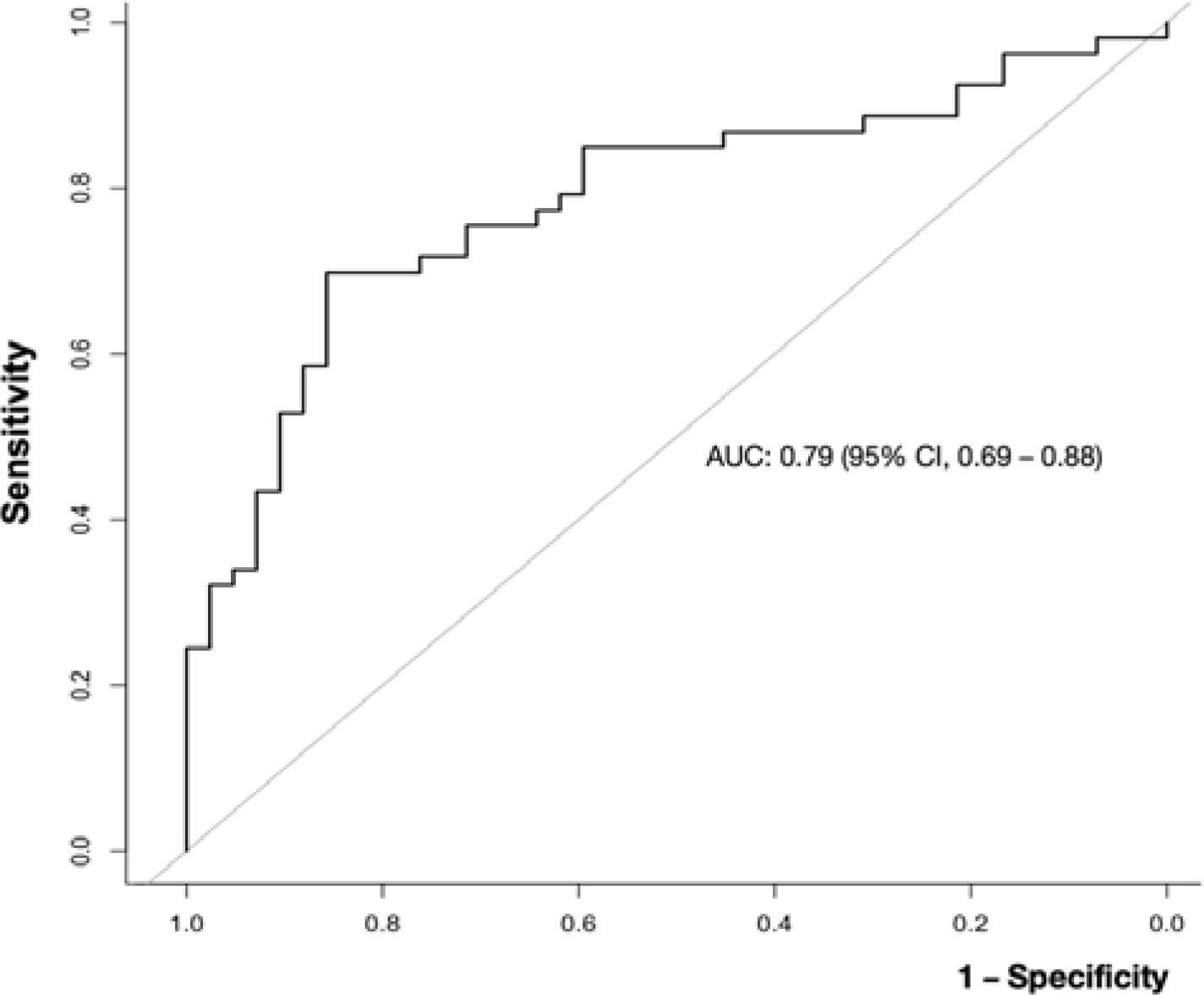
Receiver operating characteristic (ROC) curve for the ability of blood viscosity to differentiate ischemic stroke from other conditions. The area under the curve (AUC) value for blood viscosity differentiating ischemic stroke from non-stroke was 0.79 (95% confidence interval, 0.69 – 0.88).

## Discussion

To our knowledge, this is the first study to verify that patients with ischemic stroke exhibit an increase in whole blood viscosity when admitted within 24 h of the last known well time, suggesting that blood viscosity measurements can aid in differentiating ischemic stroke from other diseases. Although previous large-scale studies have highlighted the long-term effects of high blood viscosity on the risk of cardiovascular events,^17, 18^ their measurements were limited by a high degree of noise and fluctuations in blood viscosity. From this perspective, our study strengthens the evidence regarding the effects of shear stress on endothelial injury or abrasion during thrombotic events.^9^

Notably, even in patients with ischemic stroke, treatment with IV fluid was associated with a decrease in blood viscosity, which approached a level similar to that in the non-stroke group (median 23.5 vs. 14.7 *cP* at 10 *Hz*). This effect is related to the water content in normal saline or plasma solution, which are Newtonian fluids with low viscosity. In addition to highlighting the sensitivity of blood viscosity measurements, this phenomenon may provide insight into the role of hemodilution or fluid treatment in patients with stroke, the benefits of which have remained controversial for some time.^19, 20^ Studies incorporating serial measurements of blood viscosity that account for hemodilution therapy may provide more accurate results regarding the clinical value of such treatment.

Considering the sensitivity of blood viscosity measurements to external factors, immediate measurements without anticoagulant use are indispensable for clinical utilization.^13^ The direct, on-site method used in the current study is advantageous in that it provides a direct measurement of blood viscosity while allowing for precise blood volume/additives control. Moreover, POCT rheometer measurements have obtained an average of 2.3 min (median 2, IQR, 1–3) following a puncture, meaning that results were unlikely affected by spontaneous clotting, and the simplicity of the process means that no additional training is required.

This study had several limitations, including its single-center design and small sample size. In addition, blood viscosity data in healthy controls was limited, and some patients had other risk factors or diseases that may impact the diagnostic utility of blood viscosity measurements. Lastly, we still need to obtain serial blood viscosity measurements to investigate changes over time during admission, highlighting the need for more in-depth, long-term analyses.

In conclusion, this study observed the elevation of blood viscosity in ischemic stroke, which would exacerbate the shear stress along the vessel wall and promote the thrombogenic condition.

## Data Availability

The use of registered data is permitted upon request after the review.

## Acknowledgment

None

## Funding of source

This work was partly supported by the Technology Development Program (S3094866) funded by the Ministry of SMEs and Startups (MSS, Korea). The results were partly supported by “Regional Innovation Strategy (RIS)” through the National Research Foundation of Korea (NRF) funded by the Ministry of Education (MOE) (2021RIS-004).

## Disclosures

The authors, J Kang, BJ Kim, JY Kim, DY Kim, S-Y Yun, M-K Han, H-J Bae, I Park, JH Lee, YH Jo, and KH Ahn, have nothing to disclose about the study. JS Oh holds the patent for the ARS-MEDI parallel plate viscometer used in this study and is the CEO of the Rheology Solutions Co., which manufactured the viscometer.

## Authors’ contributions

— Study concept and design: J Kang
— Acquisition of data: J Kang, BJ Kim, JY Kim, DY Kim, S-Y Yun, M-K Han, H-J Bae, I Park, JH Lee, YH Jo
— Analysis and interpretation of data: J Kang, DY Kim, S-Y Yun, J-S Oh, I Park, JH Lee, YH Jo, and KH Ahn
— Drafting of the manuscript: J Kang
— All authors read and approved the final manuscript.

## Notes

### Clinical Trial

This study is a prospective observational study and not registered to the ACMJE-approved registry.

### Author Declarations

The local institutional review board of Seoul National University Bundang Hospital (Republic of Korea) approved this study. Patients or caregivers provided written informed consent for additional blood sampling and viscosity measurements.

